# Early initiation of prophylactic anticoagulation for prevention of COVID-19 mortality: a nationwide cohort study of hospitalized patients in the United States

**DOI:** 10.1101/2020.12.09.20246579

**Authors:** Christopher T Rentsch, Joshua A Beckman, Laurie Tomlinson, Walid F Gellad, Charles Alcorn, Farah Kidwai-Khan, Melissa Skanderson, Evan Brittain, Joseph T King, Yuk-Lam Ho, Svetlana Eden, Suman Kundu, Michael F Lann, Robert A Greevy, P. Michael Ho, Paul A Heidenreich, Daniel A Jacobson, Ian J Douglas, Janet P Tate, Stephen JW Evans, David Atkins, Amy C Justice, Matthew S Freiberg

## Abstract

**Importance:** Deaths among patients with coronavirus disease 2019 (COVID-19) are partially attributed to venous thromboembolism and arterial thromboses. Anticoagulants prevent thrombosis formation, possess anti-inflammatory and anti-viral properties, and may be particularly effective for treating patients with COVID-19.

**Objective:** To evaluate whether initiation of prophylactic anticoagulation within 24 hours of admission is associated with decreased risk of death among patients hospitalized with COVID-19.

**Design:** Observational cohort study.

**Setting:** Nationwide cohort of patients receiving care in the Department of Veterans Affairs, the largest integrated healthcare system in the United States.

**Participants:** All patients hospitalized with laboratory-confirmed SARS-CoV-2 infection March 1 to July 31, 2020, without a history of therapeutic anticoagulation.

**Exposures:** Prophylactic doses of subcutaneous heparin, low-molecular-weight heparin, or direct oral anticoagulants.

**Main Outcomes and Measures:** 30-day mortality. Secondary outcomes: inpatient mortality and initiating therapeutic anticoagulation.

**Results:** Of 4,297 patients hospitalized with COVID-19, 3,627 (84.4%) received prophylactic anticoagulation within 24 hours of admission. More than 99% (n=3,600) received subcutaneous heparin or enoxaparin. We observed 622 deaths within 30 days of admission, 513 among those who received prophylactic anticoagulation. Most deaths (510/622, 82%) occurred during hospitalization. In inverse probability of treatment weighted analyses, cumulative adjusted incidence of mortality at 30 days was 14.3% (95% CI 13.1-15.5) among those receiving prophylactic anticoagulation and 18.7% (95% CI 15.1-22.9) among those who did not. Compared to patients who did not receive prophylactic anticoagulation, those who did had a 27% decreased risk for 30-day mortality (HR 0.73, 95% CI 0.66-0.81). Similar associations were found for inpatient mortality and initiating therapeutic anticoagulation. Quantitative bias analysis demonstrated that results were robust to unmeasured confounding (e-value lower 95% CI 1.77). Results persisted in a number of sensitivity analyses.

**Conclusions and Relevance:** Early initiation of prophylactic anticoagulation among patients hospitalized with COVID-19 was associated with a decreased risk of mortality. These findings provide strong real-world evidence to support guidelines recommending the use of prophylactic anticoagulation as initial therapy for COVID-19 patients upon hospital admission.

## Background

Severe acute respiratory syndrome coronavirus 2 (SARS-CoV-2), the virus that causes coronavirus disease 2019 (COVID-19), continues to spread worldwide. Deaths among individuals with COVID-19 have been partially attributed to venous thromboembolism and arterial thromboses.^1,2^ In intensive care settings, prevalence of venous thromboembolism among patients with COVID-19 has been reported to be over 40%.^3^ In response, several expert organizations including the American Society of Hematology, the International Society on Thrombosis and Haemostasis, and the CHEST Guideline and Expert Panel have recommended the use of prophylactic anticoagulation for patients admitted with COVID-19 who do not have a contraindication to this therapy.^1,4,5^

The most commonly used anticoagulants in hospital settings are heparin-based. Given these drugs also possess anti-inflammatory properties,^6–8^ heparin-based therapies may be particularly effective in treating patients with COVID-19.^9^ Evaluations of the efficacy of prophylactic anticoagulation in COVID-19 patients in randomized clinical trials are underway but yet to be reported.^10^ Previous observational cohort studies have shown evidence that use of anticoagulation in COVID-19 patients was associated with decreased risk of mortality;^11,12^ however, these studies were limited in sample size or relatively small healthcare systems. Our objective was to estimate the effect of early initiation of prophylactic anticoagulation on the risk of 30-day mortality among patients hospitalized with COVID-19 in the largest integrated healthcare system in the United States.

## Methods

### Study design and population

We conducted an observational cohort study using electronic health record (EHR) data from the US Department of Veterans Affairs (VA), which comprises over 1,200 points of care nationwide including hospitals, medical centers, and community outpatient clinics. All care is recorded in an EHR with daily uploads into the VA Corporate Data Warehouse. Available data include demographics, outpatient and inpatient encounters, diagnoses, procedures, smoking and alcohol health behaviors, pharmacy dispensing records, vital signs, laboratory measures, and death information.

We included all patients hospitalized between March 1 and July 31, 2020 who had a laboratory-confirmed positive SARS-CoV-2 test result on or within 14 days prior to hospital admission. We excluded patients who had no history of care (defined as at least one outpatient or inpatient encounter in the two years prior to March 1, 2020), received therapeutic anticoagulation in the 30 days prior to hospital admission (to mitigate the effect of prevalent use of anticoagulation), received a red blood cell transfusion with 24 hours of admission (as active bleeding or severe anemia may have been a contraindication for anticoagulation), or experienced any of the primary outcomes (i.e., died or initiated therapeutic anticoagulation) within 24 hours of admission and therefore did not have equal chance to be classified as exposed in this study.

### Forms and doses of anticoagulation

We extracted inpatient pharmacy records for warfarin, intravenous heparin, low-molecular-weight heparin (LMWH; i.e., enoxaparin, fondaparinux, dalteparin), and direct oral anticoagulants (DOAC; i.e., apixaban, rivaroxaban, dabigatran). Doses and routes considered prophylactic anticoagulation are listed in **Box 1**. Any dose higher than these levels, in addition to warfarin at any dose, were considered therapeutic anticoagulation.

**Box 1.** Agents and doses of prophylactic anticoagulation

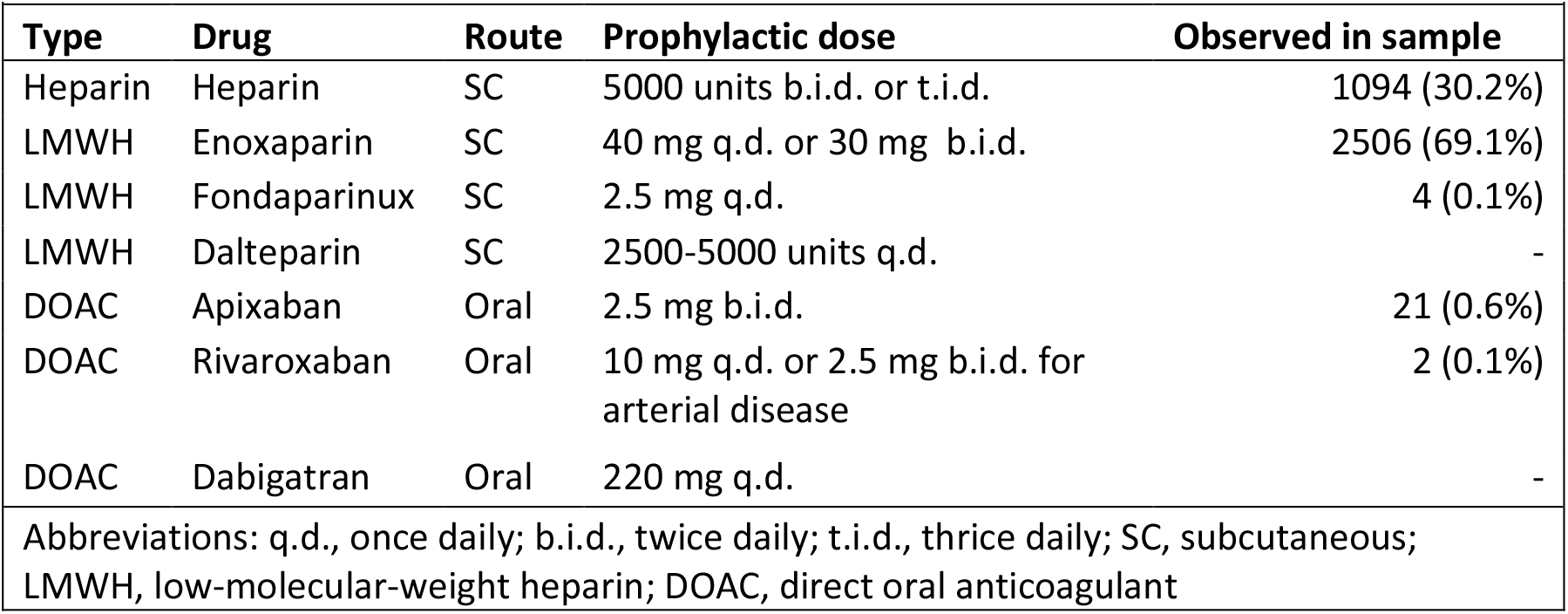

### Exposure, outcomes, and follow-up

The exposure of interest was receipt of prophylactic anticoagulation in the first 24 hours of hospitalization compared to no receipt of prophylactic anticoagulation in the same time frame. Primary outcome was mortality within 30 days of hospital admission (“30-day mortality”), which included in-hospital deaths as well as those that occurred after discharge. Secondary outcomes were inpatient mortality and initiation of therapeutic anticoagulation. Algorithms to identify thromboembolic events during hospitalizations of COVID-19 patients have yet to be validated; thus, we considered initiation of therapeutic levels of anticoagulation after the first 24 hours of admission a proxy for a thromboembolic event. For all outcomes, we followed patients from date of hospital admission until earliest of date of outcome or a maximum of 30 days.

Some VA hospitals report observation periods and admissions separately, even when a patient has not moved beds or changed providers. We combined these periods and considered a full hospitalization to begin at first presentation in a VA hospital and end when there was not a subsequent “stay” that began within 24 hours. Study design is depicted in **Figure 1**.

**Figure 1.**
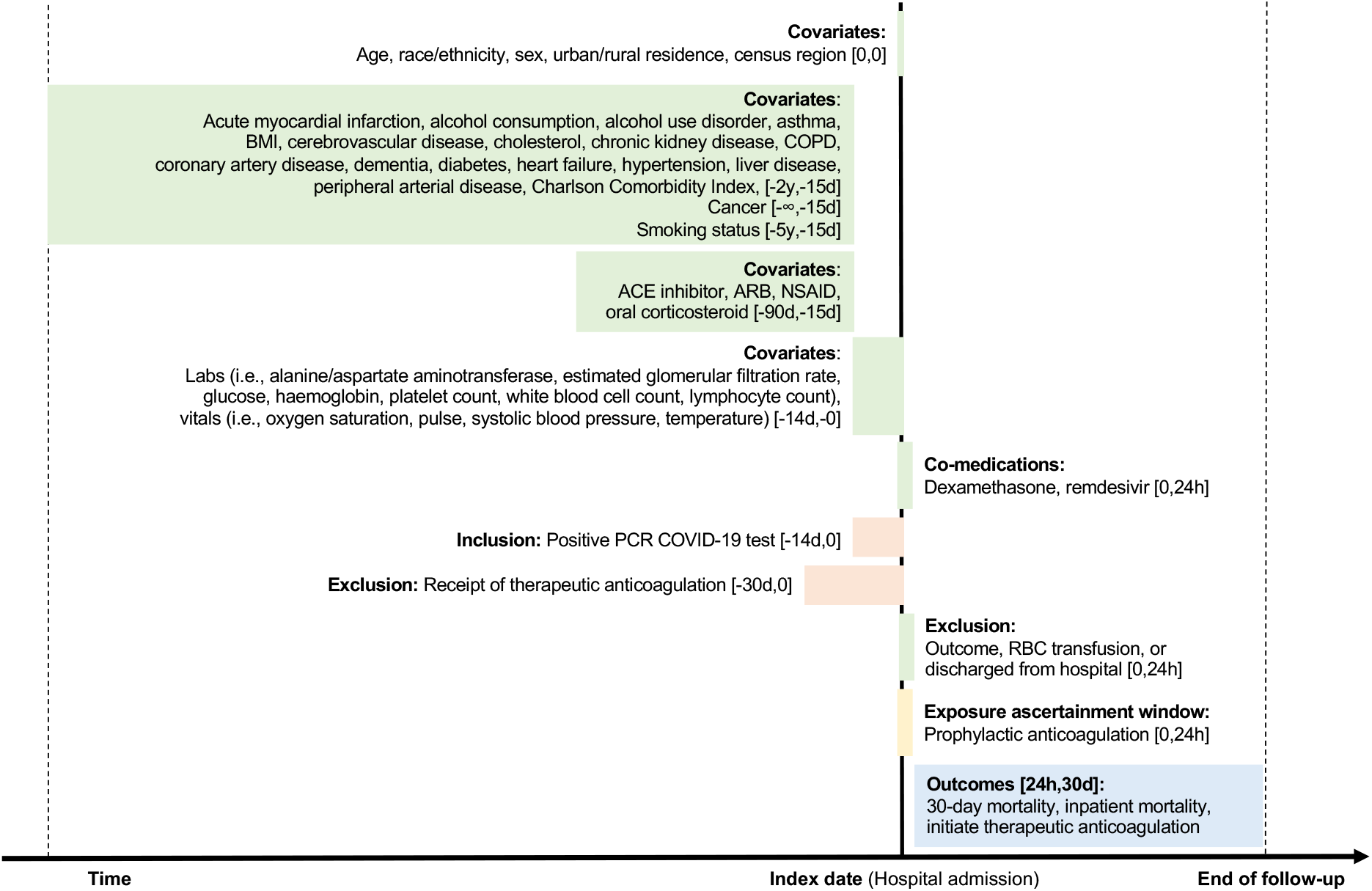
Study diagram *Abbreviations:* h, hour; d, day; m, month; y, year; BMI, body mass index; COPD, chronic obstructive pulmonary disease; ACE, angiotensin converting enzyme; ARB, angiotensin II receptor blocker; NSAID, non-steroidal anti-inflammatory drug; PCR, polymerase chain reaction; RBC, red blood cell*Notes:* End of follow-up for all outcomes was earliest of: date of outcome, a maximum of 30 days, or August 30, 2020. For the analysis of inpatient mortality and initiating therapeutic anticoagulation, we further censored patients at date of hospital discharge.

### Covariates

Potential confounders in the relationship between receipt of anticoagulation and COVID-19 mortality or thromboembolic events were identified by reviewing existing literature and through discussions with clinicians. We extracted information on age, race/ethnicity, sex, urban/rural residence, US Census region, clinical comorbidities, Charlson Comorbidity Index, and substance use. Presence of clinical comorbidities was determined by one inpatient or two outpatient diagnoses using International Classification of Diseases − 9th or 10th edition (ICD-9/10) codes in the two years prior to hospitalization, except for cancer, which was considered present if diagnosed ever prior to hospitalization. Level of alcohol consumption was calculated using the most recent Alcohol Use Disorder Identification Test - Consumption (AUDIT-C)^13^ measure within two years prior to admission. Smoking status was determined by the most frequent response in the five years prior to hospitalization.^14^

We ascertained medication history in the 15 to 90 days prior to hospitalization. We captured vital signs and laboratory measures to account for acute health status at hospital admission. Height and weight measurements closest to hospitalization within five and two years, respectively, prior to admission were used to calculate body mass index. All other vital signs and all laboratory measures utilized the value closest to hospitalization within 14 days prior to admission. Further details on covariate ascertainment windows are depicted in **Figure 1**.

To account for potential effects of co-medications with other COVID-19 treatments, we ascertained receipt of oral or intravenous dexamethasone^15^ at any dose or intravenous remdesivir^16^ at any dose within the first 24 hours of hospitalization as well as treatments received after the first 24 hours.

Covariates with the largest proportion of missing data included alanine aminotransferase (13.5%), aspartate aminotransferase (15.2%), lymphocyte count (15.0%), and total cholesterol (14.1%): all other covariates had <10% missing.

### Propensity score model

We used inverse probability of treatment (IPT) weighting to estimate the marginal treatment effect. We first modeled the probability of receiving the exposure of interest as a function of all measured covariates (apart from in-hospital treatments received after the first 24 hours so as to not use future information at baseline).^17^ Propensity scores (i.e., the predicted probability of exposure) were estimated using a multivariable logistic regression model. We included a missing category for covariates with missing data. The area under the receiver operating characteristic curve was 0.74. Each patient was weighted by the inverse probability of receiving the exposure of interest, with the goal of balancing observable characteristics between treatment groups. After IPT weighting, the distribution of propensity scores between the treatment groups overlapped nearly perfectly (**eFigure 1**). We calculated absolute standardized mean differences between treatment groups and considered ≤0.2 as balanced,18 although the vast majority were ≤0.1 (**eFigure 2**). Thus, the weighting produced treatment groups that were considered well balanced.

**Figure 2.**
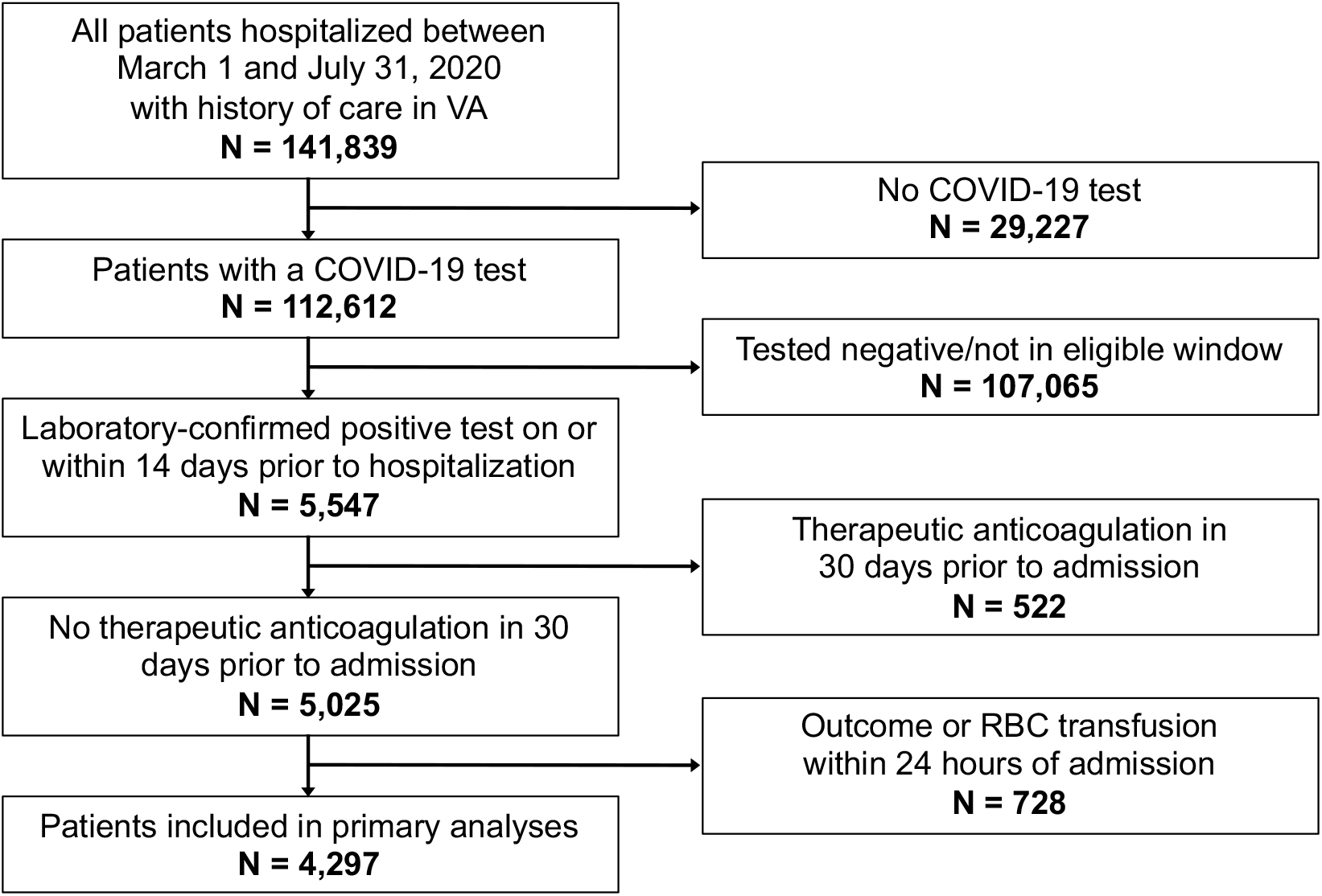
Flow chart *Abbreviations:* VA, US Department of Veterans Affairs; COVID-19, coronavirus disease 2019; RBC, red blood cell

### Statistical methods

Covariates were summarized using descriptive statistics, stratified by treatment group. We generated IPT-weighted Kaplan Meier plots. We used Cox regression models with days since hospital admission as the timescale to estimate IPT-weighted hazard ratios (HRs) and 95% confidence intervals (CIs) for the effect of early initiation of prophylactic doses of anticoagulation on 30-day mortality, inpatient mortality, and initiating therapeutic anticoagulation. Both secondary outcomes occur during hospitalization, wherein discharge from hospital was considered a competing event. If patients were censored at discharge, absolute risks derived from Kaplan-Meier analyses would be overestimated.^19,20^ We therefore displayed cumulative adjusted incidence rates by treatment group treating discharge as a competing event (i.e., no censoring at date of discharge). We nonetheless chose to display cause-specific rather than sub-distribution hazard ratios as our question was etiological in nature.^21^ These cause-specific hazard ratios were interpreted as the effect of prophylactic anticoagulation on each of the outcomes irrespective of its effect on discharge. Proportional hazards were checked by examining the complementary log-log (or the log of negative log) of estimated survivor functions for exposed and unexposed groups versus the log of survival time. There was no evidence of proportional hazards violations.

### Sensitivity analyses

For each outcome, we used quantitative bias analysis to calculate an e-value, which demonstrates the strength of association between an unmeasured confounder and exposure or outcome, conditional on measured covariates, that would be necessary to fully explain observed effects.^22^ To assess for undue effects from outliers with very high or very low estimated propensity of treatment, we capped propensity score distributions at the 1^st^/99^th^ and again at the 5^th^/95^th^ percentiles. To account for potentially biased estimation of standard errors or influence from very high or very low weights, we performed sensitivity analyses using combinations of robust standard error estimation^23^ and stabilized weighting.^24^ We re-ran the primary analyses extending the exposure ascertainment window from 24 to 48 hours. Given the low frequency of DOAC use in the cohort, we re-ran analyses excluding DOACs from the exposure definition. In post-hoc analyses, we assessed the effect of prophylactic anticoagulation separately by the two most commonly prescribed drugs in the cohort, heparin and enoxaparin. We used Microsoft SQL Server Management Studio v17.4 for data management and SAS version 9.4 (SAS Institute, Cary, NC, US) and Stata 16 MP for statistical analyses.

### Ethics

This study was approved by the institutional review boards of VA Connecticut Healthcare System and Yale University. It has been granted a waiver of informed consent and is Health Insurance Portability and Accountability Act compliant. This study is reported as per the Strengthening the Reporting of Observational Studies in Epidemiology (STROBE) and REporting of studies Conducted using Observational Routinely collected health Data for pharmacoepidemiology (RECORD-PE) guidelines (**Supplementary Appendix**).

## Results

### Patient characteristics

We identified 4,297 patients hospitalized with COVID-19 between March 1 and July 31, 2020 who were included in this analysis (**Figure 2**). Median age in the cohort was 68 years (interquartile range [IQR] 58-75); most were non-Hispanic Black (n=1,940, n=45.1%), non-Hispanic White (n=1,603, 37.3%), or Hispanic (n=506, 11.8%). The majority were male (n=4,015, 93.4%), geographically located in the South (n=2,017, 46.9%), and lived in an urban area (n=3,768, 87.7%) (**Table 1**). By month, most patients were hospitalized in July (n=1,401, 32.6%).

**Table 1.**
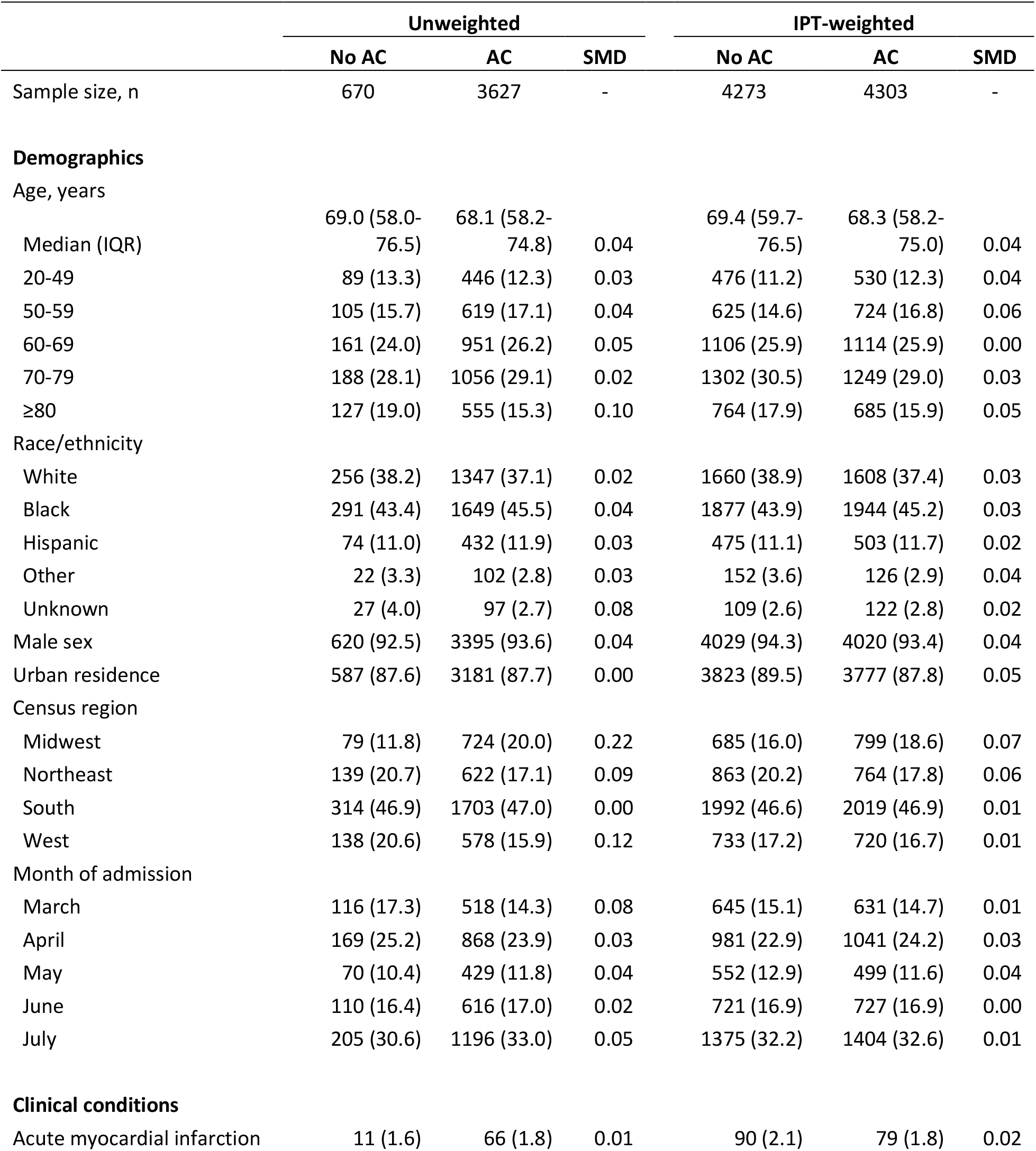
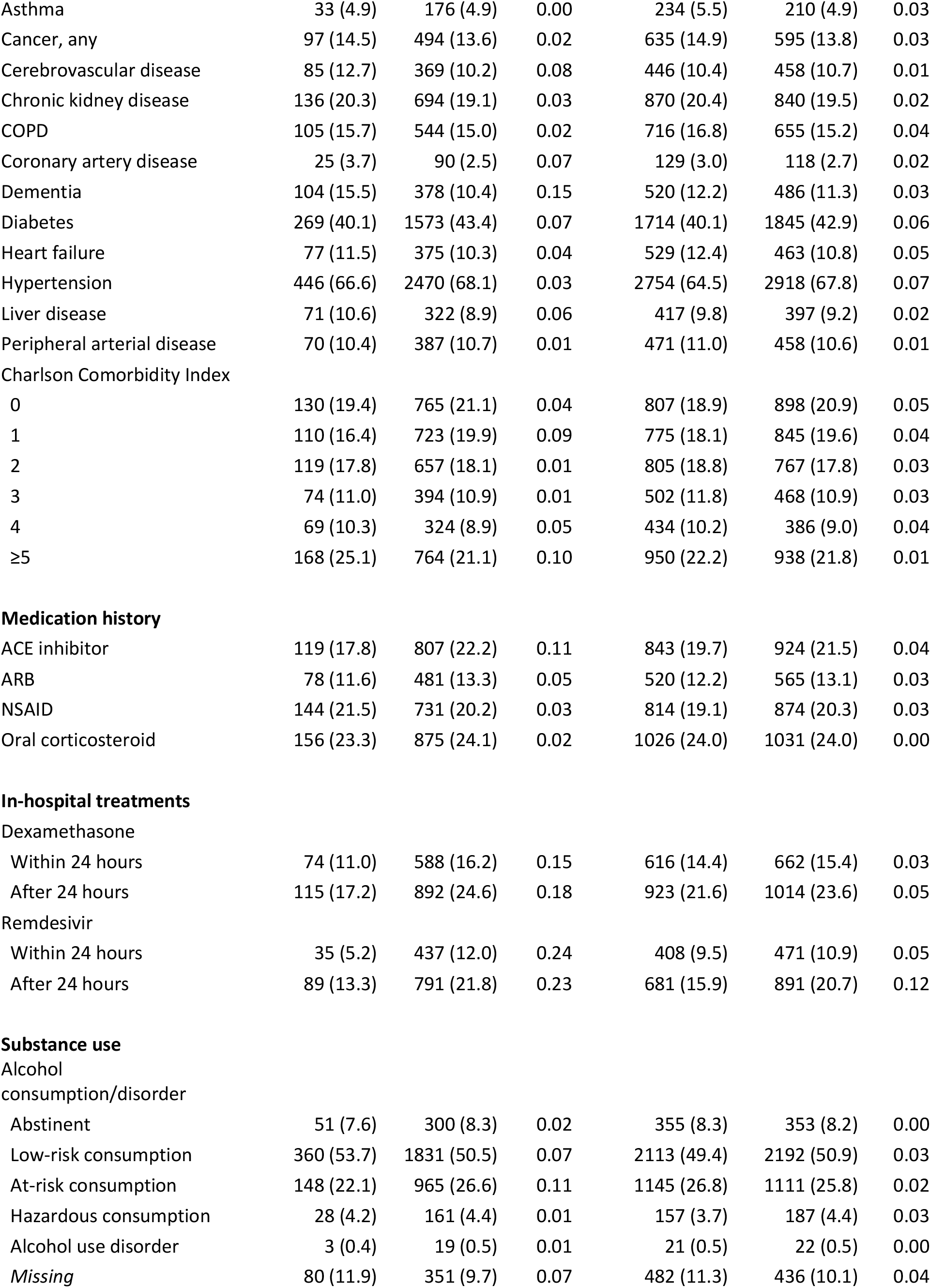
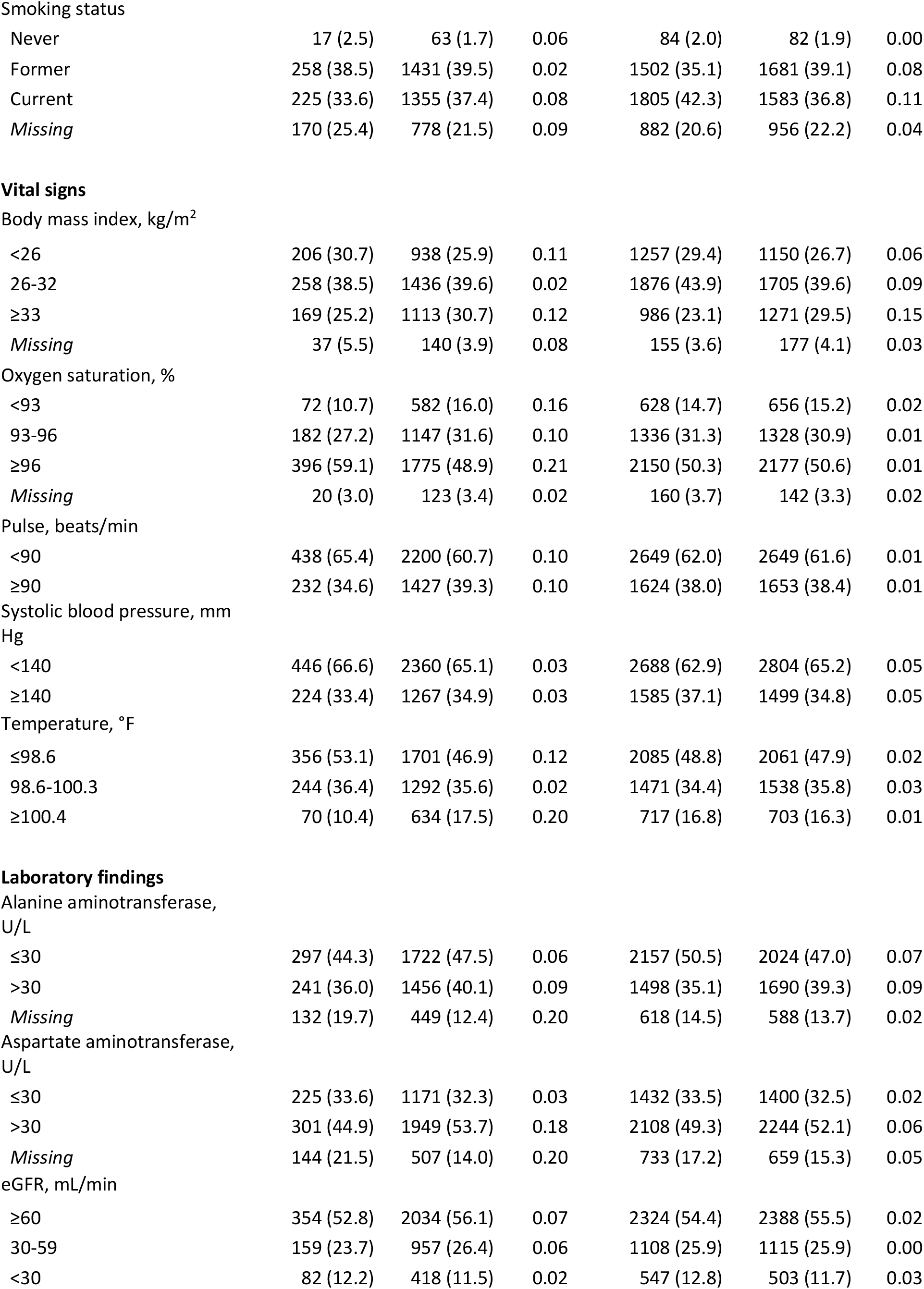
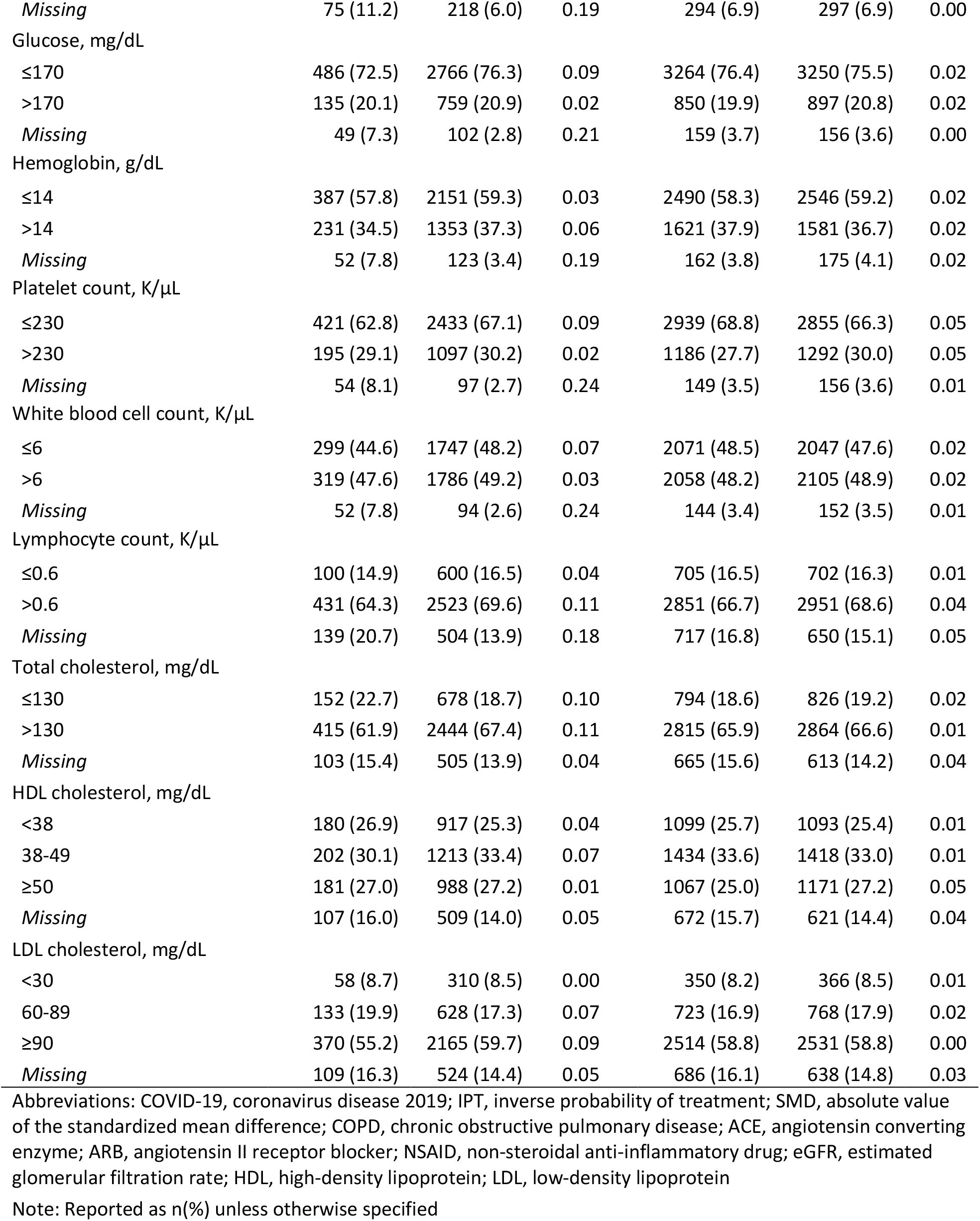
Demographic and clinical characteristics of 4,297 hospitalized patients with COVID-19 by exposure to prophylactic dose of anticoagulation (AC) within 24 hours of admission, before and after weighting

|  | Unweighted |  |  | IPT-weighted |  |  |
| --- | --- | --- | --- | --- | --- | --- |
|  | No AC | AC | SMD | No AC | AC | SMD |
| Sample size, n | 670 | 3627 | - | 4273 | 4303 | - |
| **Demographics** |  |  |  |  |  |  |
| Age, years |  |  |  |  |  |  |
| Median (IQR) | 69.0 (58.0-76.5) | 68.1 (58.2-74.8) | 0.04 | 69.4 (59.7-76.5) | 68.3 (58.2-75.0) | 0.04 |
| 20-49 | 89 (13.3) | 446 (12.3) | 0.03 | 476 (11.2) | 530 (12.3) | 0.04 |
| 50-59 | 105 (15.7) | 619 (17.1) | 0.04 | 625 (14.6) | 724 (16.8) | 0.06 |
| 60-69 | 161 (24.0) | 951 (26.2) | 0.05 | 1106 (25.9) | 1114 (25.9) | 0.00 |
| 70-79 | 188 (28.1) | 1056 (29.1) | 0.02 | 1302 (30.5) | 1249 (29.0) | 0.03 |
| ≥80 | 127 (19.0) | 555 (15.3) | 0.10 | 764 (17.9) | 685 (15.9) | 0.05 |
| Race/ethnicity |  |  |  |  |  |  |
| White | 256 (38.2) | 1347 (37.1) | 0.02 | 1660 (38.9) | 1608 (37.4) | 0.03 |
| Black | 291 (43.4) | 1649 (45.5) | 0.04 | 1877 (43.9) | 1944 (45.2) | 0.03 |
| Hispanic | 74 (11.0) | 432 (11.9) | 0.03 | 475 (11.1) | 503 (11.7) | 0.02 |
| Other | 22 (3.3) | 102 (2.8) | 0.03 | 152 (3.6) | 126 (2.9) | 0.04 |
| Unknown | 27 (4.0) | 97 (2.7) | 0.08 | 109 (2.6) | 122 (2.8) | 0.02 |
| Male sex | 620 (92.5) | 3395 (93.6) | 0.04 | 4029 (94.3) | 4020 (93.4) | 0.04 |
| Urban residence | 587 (87.6) | 3181 (87.7) | 0.00 | 3823 (89.5) | 3777 (87.8) | 0.05 |
| Census region |  |  |  |  |  |  |
| Midwest | 79 (11.8) | 724 (20.0) | 0.22 | 685 (16.0) | 799 (18.6) | 0.07 |
| Northeast | 139 (20.7) | 622 (17.1) | 0.09 | 863 (20.2) | 764 (17.8) | 0.06 |
| South | 314 (46.9) | 1703 (47.0) | 0.00 | 1992 (46.6) | 2019 (46.9) | 0.01 |
| West | 138 (20.6) | 578 (15.9) | 0.12 | 733 (17.2) | 720 (16.7) | 0.01 |
| Month of admission |  |  |  |  |  |  |
| March | 116 (17.3) | 518 (14.3) | 0.08 | 645 (15.1) | 631 (14.7) | 0.01 |
| April | 169 (25.2) | 868 (23.9) | 0.03 | 981 (22.9) | 1041 (24.2) | 0.03 |
| May | 70 (10.4) | 429 (11.8) | 0.04 | 552 (12.9) | 499 (11.6) | 0.04 |
| June | 110 (16.4) | 616 (17.0) | 0.02 | 721 (16.9) | 727 (16.9) | 0.00 |
| July | 205 (30.6) | 1196 (33.0) | 0.05 | 1375 (32.2) | 1404 (32.6) | 0.01 |
| **Clinical conditions** |  |  |  |  |  |  |
| Acute myocardial infarction | 11 (1.6) | 66 (1.8) | 0.01 | 90 (2.1) | 79 (1.8) | 0.02 |
| Asthma | 33 (4.9) | 176 (4.9) | 0.00 | 234 (5.5) | 210 (4.9) | 0.03 |
| Cancer, any | 97 (14.5) | 494 (13.6) | 0.02 | 635 (14.9) | 595 (13.8) | 0.03 |
| Cerebrovascular disease | 85 (12.7) | 369 (10.2) | 0.08 | 446 (10.4) | 458 (10.7) | 0.01 |
| Chronic kidney disease | 136 (20.3) | 694 (19.1) | 0.03 | 870 (20.4) | 840 (19.5) | 0.02 |
| COPD | 105 (15.7) | 544 (15.0) | 0.02 | 716 (16.8) | 655 (15.2) | 0.04 |
| Coronary artery disease | 25 (3.7) | 90 (2.5) | 0.07 | 129 (3.0) | 118 (2.7) | 0.02 |
| Dementia | 104 (15.5) | 378 (10.4) | 0.15 | 520 (12.2) | 486 (11.3) | 0.03 |
| Diabetes | 269 (40.1) | 1573 (43.4) | 0.07 | 1714 (40.1) | 1845 (42.9) | 0.06 |
| Heart failure | 77 (11.5) | 375 (10.3) | 0.04 | 529 (12.4) | 463 (10.8) | 0.05 |
| Hypertension | 446 (66.6) | 2470 (68.1) | 0.03 | 2754 (64.5) | 2918 (67.8) | 0.07 |
| Liver disease | 71 (10.6) | 322 (8.9) | 0.06 | 417 (9.8) | 397 (9.2) | 0.02 |
| Peripheral arterial disease | 70 (10.4) | 387 (10.7) | 0.01 | 471 (11.0) | 458 (10.6) | 0.01 |
| Charlson Comorbidity Index |  |  |  |  |  |  |
| 0 | 130 (19.4) | 765 (21.1) | 0.04 | 807 (18.9) | 898 (20.9) | 0.05 |
| 1 | 110 (16.4) | 723 (19.9) | 0.09 | 775 (18.1) | 845 (19.6) | 0.04 |
| 2 | 119 (17.8) | 657 (18.1) | 0.01 | 805 (18.8) | 767 (17.8) | 0.03 |
| 3 | 74 (11.0) | 394 (10.9) | 0.01 | 502 (11.8) | 468 (10.9) | 0.03 |
| 4 | 69 (10.3) | 324 (8.9) | 0.05 | 434 (10.2) | 386 (9.0) | 0.04 |
| ≥5 | 168 (25.1) | 764 (21.1) | 0.10 | 950 (22.2) | 938 (21.8) | 0.01 |
| **Medication history** |  |  |  |  |  |  |
| ACE inhibitor | 119 (17.8) | 807 (22.2) | 0.11 | 843 (19.7) | 924 (21.5) | 0.04 |
| ARB | 78 (11.6) | 481 (13.3) | 0.05 | 520 (12.2) | 565 (13.1) | 0.03 |
| NSAID | 144 (21.5) | 731 (20.2) | 0.03 | 814 (19.1) | 874 (20.3) | 0.03 |
| Oral corticosteroid | 156 (23.3) | 875 (24.1) | 0.02 | 1026 (24.0) | 1031 (24.0) | 0.00 |
| **In-hospital treatments** |  |  |  |  |  |  |
| Dexamethasone |  |  |  |  |  |  |
| Within 24 hours | 74 (11.0) | 588 (16.2) | 0.15 | 616 (14.4) | 662 (15.4) | 0.03 |
| After 24 hours | 115 (17.2) | 892 (24.6) | 0.18 | 923 (21.6) | 1014 (23.6) | 0.05 |
| Remdesivir |  |  |  |  |  |  |
| Within 24 hours | 35 (5.2) | 437 (12.0) | 0.24 | 408 (9.5) | 471 (10.9) | 0.05 |
| After 24 hours | 89 (13.3) | 791 (21.8) | 0.23 | 681 (15.9) | 891 (20.7) | 0.12 |
| **Substance use** |  |  |  |  |  |  |
| Alcohol consumption/disorder |  |  |  |  |  |  |
| Abstinent | 51 (7.6) | 300 (8.3) | 0.02 | 355 (8.3) | 353 (8.2) | 0.00 |
| Low-risk consumption | 360 (53.7) | 1831 (50.5) | 0.07 | 2113 (49.4) | 2192 (50.9) | 0.03 |
| At-risk consumption | 148 (22.1) | 965 (26.6) | 0.11 | 1145 (26.8) | 1111 (25.8) | 0.02 |
| Hazardous consumption | 28 (4.2) | 161 (4.4) | 0.01 | 157 (3.7) | 187 (4.4) | 0.03 |
| Alcohol use disorder | 3 (0.4) | 19 (0.5) | 0.01 | 21 (0.5) | 22 (0.5) | 0.00 |
| Missing | 80 (11.9) | 351 (9.7) | 0.07 | 482 (11.3) | 436 (10.1) | 0.04 |
| Smoking status |  |  |  |  |  |  |
| Never | 17 (2.5) | 63 (1.7) | 0.06 | 84 (2.0) | 82 (1.9) | 0.00 |
| Former | 258 (38.5) | 1431 (39.5) | 0.02 | 1502 (35.1) | 1681 (39.1) | 0.08 |
| Current | 225 (33.6) | 1355 (37.4) | 0.08 | 1805 (42.3) | 1583 (36.8) | 0.11 |
| Missing | 170 (25.4) | 778 (21.5) | 0.09 | 882 (20.6) | 956 (22.2) | 0.04 |
| **Vital signs** |  |  |  |  |  |  |
| Body mass index, kg/m<sup>2</sup> |  |  |  |  |  |  |
| <26 | 206 (30.7) | 938 (25.9) | 0.11 | 1257 (29.4) | 1150 (26.7) | 0.06 |
| 26-32 | 258 (38.5) | 1436 (39.6) | 0.02 | 1876 (43.9) | 1705 (39.6) | 0.09 |
| ≥33 | 169 (25.2) | 1113 (30.7) | 0.12 | 986 (23.1) | 1271 (29.5) | 0.15 |
| Missing | 37 (5.5) | 140 (3.9) | 0.08 | 155 (3.6) | 177 (4.1) | 0.03 |
| Oxygen saturation, % |  |  |  |  |  |  |
| <93 | 72 (10.7) | 582 (16.0) | 0.16 | 628 (14.7) | 656 (15.2) | 0.02 |
| 93-96 | 182 (27.2) | 1147 (31.6) | 0.10 | 1336 (31.3) | 1328 (30.9) | 0.01 |
| ≥96 | 396 (59.1) | 1775 (48.9) | 0.21 | 2150 (50.3) | 2177 (50.6) | 0.01 |
| Missing | 20 (3.0) | 123 (3.4) | 0.02 | 160 (3.7) | 142 (3.3) | 0.02 |
| Pulse, beats/min |  |  |  |  |  |  |
| <90 | 438 (65.4) | 2200 (60.7) | 0.10 | 2649 (62.0) | 2649 (61.6) | 0.01 |
| ≥90 | 232 (34.6) | 1427 (39.3) | 0.10 | 1624 (38.0) | 1653 (38.4) | 0.01 |
| Systolic blood pressure, mm Hg |  |  |  |  |  |  |
| <140 | 446 (66.6) | 2360 (65.1) | 0.03 | 2688 (62.9) | 2804 (65.2) | 0.05 |
| ≥140 | 224 (33.4) | 1267 (34.9) | 0.03 | 1585 (37.1) | 1499 (34.8) | 0.05 |
| Temperature, °F |  |  |  |  |  |  |
| ≤98.6 | 356 (53.1) | 1701 (46.9) | 0.12 | 2085 (48.8) | 2061 (47.9) | 0.02 |
| 98.6-100.3 | 244 (36.4) | 1292 (35.6) | 0.02 | 1471 (34.4) | 1538 (35.8) | 0.03 |
| ≥100.4 | 70 (10.4) | 634 (17.5) | 0.20 | 717 (16.8) | 703 (16.3) | 0.01 |
| **Laboratory findings** |  |  |  |  |  |  |
| Alanine aminotransferase, U/L |  |  |  |  |  |  |
| ≤30 | 297 (44.3) | 1722 (47.5) | 0.06 | 2157 (50.5) | 2024 (47.0) | 0.07 |
| >30 | 241 (36.0) | 1456 (40.1) | 0.09 | 1498 (35.1) | 1690 (39.3) | 0.09 |
| Missing | 132 (19.7) | 449 (12.4) | 0.20 | 618 (14.5) | 588 (13.7) | 0.02 |
| Aspartate aminotransferase, U/L |  |  |  |  |  |  |
| ≤30 | 225 (33.6) | 1171 (32.3) | 0.03 | 1432 (33.5) | 1400 (32.5) | 0.02 |
| >30 | 301 (44.9) | 1949 (53.7) | 0.18 | 2108 (49.3) | 2244 (52.1) | 0.06 |
| Missing | 144 (21.5) | 507 (14.0) | 0.20 | 733 (17.2) | 659 (15.3) | 0.05 |
| eGFR, mL/min |  |  |  |  |  |  |
| ≥60 | 354 (52.8) | 2034 (56.1) | 0.07 | 2324 (54.4) | 2388 (55.5) | 0.02 |
| 30-59 | 159 (23.7) | 957 (26.4) | 0.06 | 1108 (25.9) | 1115 (25.9) | 0.00 |
| <30 | 82 (12.2) | 418 (11.5) | 0.02 | 547 (12.8) | 503 (11.7) | 0.03 |
| *Missing* | 75 (11.2) | 218 (6.0) | 0.19 | 294 (6.9) | 297 (6.9) | 0.00 |
| Glucose, mg/dL |  |  |  |  |  |  |
| ≤170 | 486 (72.5) | 2766 (76.3) | 0.09 | 3264 (76.4) | 3250 (75.5) | 0.02 |
| >170 | 135 (20.1) | 759 (20.9) | 0.02 | 850 (19.9) | 897 (20.8) | 0.02 |
| *Missing* | 49 (7.3) | 102 (2.8) | 0.21 | 159 (3.7) | 156 (3.6) | 0.00 |
| Hemoglobin, g/dL |  |  |  |  |  |  |
| ≤14 | 387 (57.8) | 2151 (59.3) | 0.03 | 2490 (58.3) | 2546 (59.2) | 0.02 |
| >14 | 231 (34.5) | 1353 (37.3) | 0.06 | 1621 (37.9) | 1581 (36.7) | 0.02 |
| *Missing* | 52 (7.8) | 123 (3.4) | 0.19 | 162 (3.8) | 175 (4.1) | 0.02 |
| Platelet count, K/μL |  |  |  |  |  |  |
| ≤230 | 421 (62.8) | 2433 (67.1) | 0.09 | 2939 (68.8) | 2855 (66.3) | 0.05 |
| >230 | 195 (29.1) | 1097 (30.2) | 0.02 | 1186 (27.7) | 1292 (30.0) | 0.05 |
| *Missing* | 54 (8.1) | 97 (2.7) | 0.24 | 149 (3.5) | 156 (3.6) | 0.01 |
| White blood cell count, K/μL |  |  |  |  |  |  |
| ≤6 | 299 (44.6) | 1747 (48.2) | 0.07 | 2071 (48.5) | 2047 (47.6) | 0.02 |
| >6 | 319 (47.6) | 1786 (49.2) | 0.03 | 2058 (48.2) | 2105 (48.9) | 0.02 |
| *Missing* | 52 (7.8) | 94 (2.6) | 0.24 | 144 (3.4) | 152 (3.5) | 0.01 |
| Lymphocyte count, K/μL |  |  |  |  |  |  |
| ≤0.6 | 100 (14.9) | 600 (16.5) | 0.04 | 705 (16.5) | 702 (16.3) | 0.01 |
| >0.6 | 431 (64.3) | 2523 (69.6) | 0.11 | 2851 (66.7) | 2951 (68.6) | 0.04 |
| *Missing* | 139 (20.7) | 504 (13.9) | 0.18 | 717 (16.8) | 650 (15.1) | 0.05 |
| Total cholesterol, mg/dL |  |  |  |  |  |  |
| ≤130 | 152 (22.7) | 678 (18.7) | 0.10 | 794 (18.6) | 826 (19.2) | 0.02 |
| >130 | 415 (61.9) | 2444 (67.4) | 0.11 | 2815 (65.9) | 2864 (66.6) | 0.01 |
| *Missing* | 103 (15.4) | 505 (13.9) | 0.04 | 665 (15.6) | 613 (14.2) | 0.04 |
| HDL cholesterol, mg/dL |  |  |  |  |  |  |
| <38 | 180 (26.9) | 917 (25.3) | 0.04 | 1099 (25.7) | 1093 (25.4) | 0.01 |
| 38-49 | 202 (30.1) | 1213 (33.4) | 0.07 | 1434 (33.6) | 1418 (33.0) | 0.01 |
| ≥50 | 181 (27.0) | 988 (27.2) | 0.01 | 1067 (25.0) | 1171 (27.2) | 0.05 |
| *Missing* | 107 (16.0) | 509 (14.0) | 0.05 | 672 (15.7) | 621 (14.4) | 0.04 |
| LDL cholesterol, mg/dL |  |  |  |  |  |  |
| <30 | 58 (8.7) | 310 (8.5) | 0.00 | 350 (8.2) | 366 (8.5) | 0.01 |
| 60-89 | 133 (19.9) | 628 (17.3) | 0.07 | 723 (16.9) | 768 (17.9) | 0.02 |
| ≥90 | 370 (55.2) | 2165 (59.7) | 0.09 | 2514 (58.8) | 2531 (58.8) | 0.00 |
| *Missing* | 109 (16.3) | 524 (14.4) | 0.05 | 686 (16.1) | 638 (14.8) | 0.03 |

Abbreviations: COVID-19, coronavirus disease 2019; IPT, inverse probability of treatment; SMD, absolute value of the standardized mean difference; COPD, chronic obstructive pulmonary disease; ACE, angiotensin converting enzyme; ARB, angiotensin II receptor blocker; NSAID, non-steroidal anti-inflammatory drug; eGFR, estimated glomerular filtration rate; HDL, high-density lipoprotein; LDL, low-density lipoprotein
Note: Reported as n(%) unless otherwise specified

In this cohort, 3,627 (84.4%) patients received prophylactic anticoagulation within the first 24 hours of hospital admission. Among those who received prophylactic anticoagulation, the most common drugs were heparin-based: either subcutaneous heparin (n=1,094, 30.2%) or enoxaparin (n=2,506, 69.1%; **Box 1 in the Supplementary Appendix**). At hospital presentation, the group of patients who received prophylactic anticoagulation, compared to the group of patients who did not, had a higher proportion with an oxygen saturation level less than 93% (Table 1; 16.0% versus 10.3%), heart rate at 90 beats per minute or above (39.2% versus 34.5%), and temperature of 100.4 degrees Fahrenheit or above (17.5% versus 10.6%). In contrast, the burden of prevalent comorbid disease (Charlson Comorbidity Index ≥5) was lower among those who received prophylactic anticoagulation as compared to those that did not (21.1% versus 25.1%, respectively). Co-medication with other COVID-19 treatments within the first 24 hours of admission were more common among those who received prophylactic anticoagulation compared to those who did not (16.2% versus 11.0% for dexamethasone; 12.0% versus 5.2% for remdesivir). However, after IPT-weighting, differences were minimized between the two treatment groups (all standardized mean differences ≤0.2 with vast majority ≤0.1; **Table 1**).

### Absolute and relative risks

There were 622 deaths (622/4297, 14.5%) that occurred within 30 days of hospital admission, 513 among those who received prophylactic anticoagulation (**Table 2**). Most deaths (510/622, 82%) occurred during hospitalization. In IPT-weighted analyses, cumulative adjusted incidence of mortality at 30 days was 14.3% (95% CI 13.1-15.5) and 18.7% (95% CI 15.1-22.9) for patients receiving and not receiving prophylactic anticoagulation, respectively (**Table 2**). Receipt of prophylactic anticoagulation was associated with a 27% decreased risk of death over the first 30 days (HR 0.73, 95% CI 0.66-0.81; **Figure 3**) compared to not receiving prophylactic anticoagulation. Similar associations were found for inpatient mortality (HR 0.69, 95% CI 0.61-0.77) and initiating therapeutic anticoagulation (HR 0.81, 95% CI 0.73-0.90).

**Table 2.** Absolute and relative risks associated with exposure to prophylactic doses of anticoagulation in the first 24 hours of hospitalization

| Outcome | N | No. events | Unweighted | IPT-weighted |  |
| --- | --- | --- | --- | --- | --- |
|  |  |  | HR (95% CI) | Cumulative incidence (95% CI) | HR (95% CI) |
| 30-day mortality |  |  |  |  |  |
| Exposed | 3627 | 513 | 0.85 (0.69-1.05) | 14.3 (13.1-15.5) | 0.73 (0.66-0.81) |
| Unexposed | 670 | 109 | ref | 18.7 (15.1-22.9) | ref |
| Inpatient mortality |  |  |  |  |  |
| Exposed | 3627 | 418 | 0.82 (0.66-1.03) | 11.7 (10.7-12.8) | 0.69 (0.61-0.77) |
| Unexposed | 670 | 92 | ref | 16.4 (13.0-20.5) | ref |
| Initiate therapeutic anticoagulation |  |  |  |  |  |
| Exposed | 3627 | 573 | 1.14 (0.91-1.42) | 15.6 (14.4-16.8) | 0.81 (0.73-0.90) |
| Unexposed | 670 | 92 | ref | 18.8 (15.2-23.1) | ref |

Abbreviations: PY, person-years; HR, hazard ratio; CI, confidence interval; IPT, inverse probability of treatment

**Figure 3.**
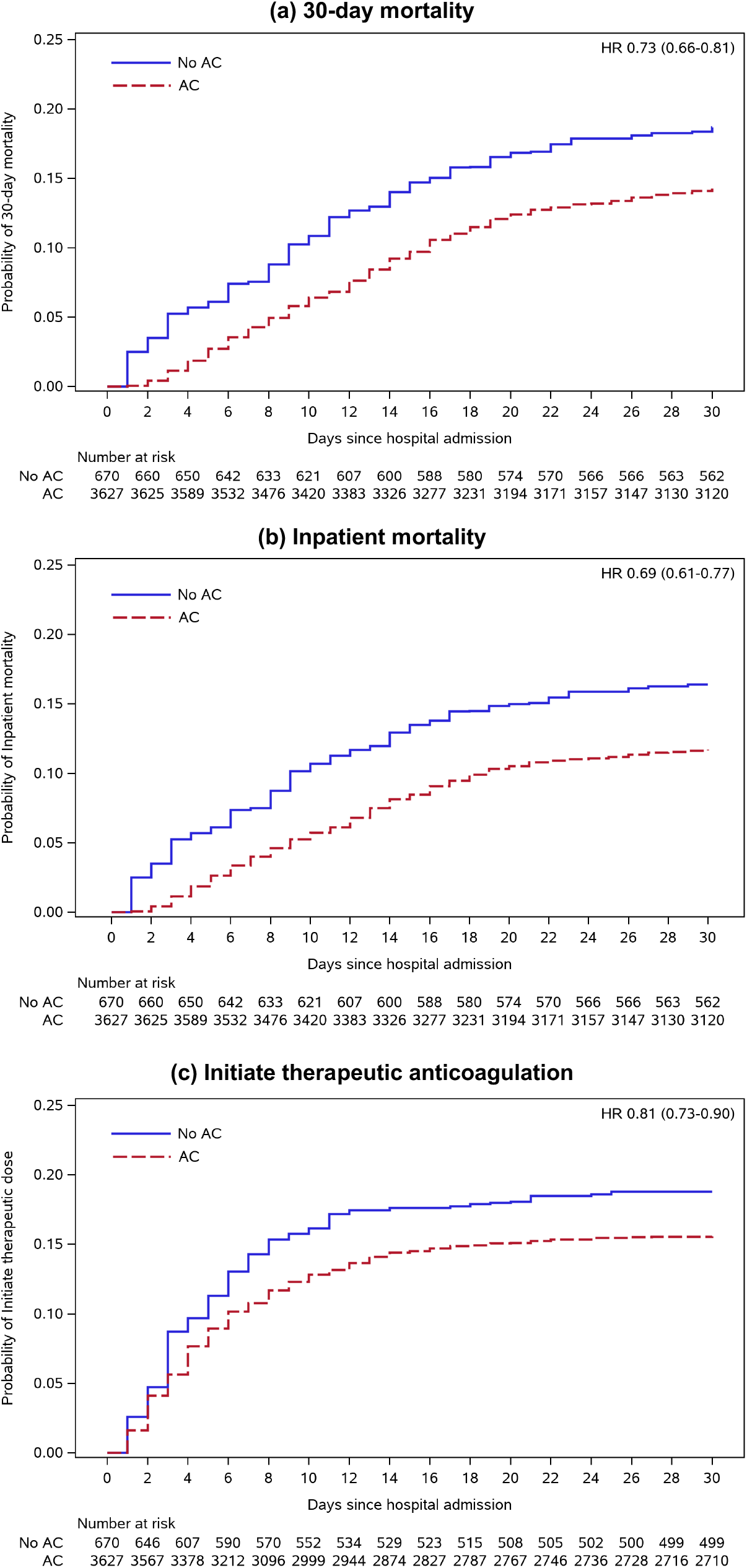
Inverse probability treatment-weighted Kaplan-Meier plots *Abbreviations:* AC, prophylactic anticoagulation

### Sensitivity analyses

Quantitative bias analysis demonstrated that an unmeasured confounder would need to be strongly associated with receipt of prophylactic anticoagulation and each outcome to explain the observed associations: e-value lower 95% CI was 1.77 for 30-day mortality, 1.92 for inpatient mortality, and 1.46 for initiating therapeutic anticoagulation (**eFigure 3**). Results were robust to capping propensity scores (**eTable 1**), using stabilized weighting and robust variance estimation (**eTable 2**), extending the exposure window from 24 to 48 hours (**eTable 3**), and excluding DOACs from the exposure definition (**eTable 4**). In post-hoc analyses, the effect of prophylactic anticoagulation on 30-day mortality was similar when stratified by whether patients received subcutaneous heparin (HR 0.73, 95% CI 0.64-0.84) or enoxaparin (HR 0.78, 95% CI 0.68-0.89; **eTable 5**).

## Discussion

### Key findings

In a nationwide cohort of 4,297 patients hospitalized with COVID-19 in the largest integrated healthcare system in the United States, initiation of predominantly heparin-based prophylactic anticoagulation within the first 24 hours of admission was associated with a relative risk reduction of 30-day mortality as high as 34% and an absolute risk reduction of 4.5% in the context of an absolute risk of 18.7% among patients who did not receive prophylactic anticoagulation. These results persisted in sensitivity analyses. We observed similar protective effects for secondary outcomes including inpatient mortality and initiation of therapeutic anticoagulation - a proxy for the development of a thromboembolic event.

### Comparison with other evidence

Previous studies investigating the role of anticoagulation among patients with COVID-19 have had varied results, but none have reported harm.^11,12,25–32^ Variations in reported associations likely derive from different exposure definitions of anticoagulation, both form and dose. Additionally, different patient populations (e.g., disease-specific cohorts), comparator groups, and inclusion/exclusion criteria were used. One of the largest observational studies to date by Nadkarni et al reported that both prophylactic and therapeutic anticoagulation were associated with a reduction in inpatient mortality by up to 55% compared to those not receiving anticoagulation across five New York City hospitals.^11^ However, the study allowed patients to switch exposure groups during follow-up without comprehensively accounting for time-updated confounding by indication that may have impacted results.

Our study was designed to emulate a hypothetical clinical trial in which we excluded prevalent users of anticoagulation, balanced covariate distribution at hospital admission (analogous to randomization) and ascertained exposure within 24 hours of admission. We conducted an intention-to-treat analysis since the data required to account for time-updated exposures and confounders (e.g., marginal structural modeling) may not be available within acute hospitalizations. For example, many hospital systems report all diagnoses that occur during a given hospitalization at discharge. Therefore, we could not determine if initiation of prophylactic anticoagulation later in hospitalization confers benefit. There are multiple clinical trials in progress to determine dosing and timing for anticoagulation during the clinical course of COVID-19.^10^ Until clinical trial data are available, our results provide strong evidence for the use of prophylactic anticoagulation as initial therapy for COVID-19 patients upon hospital admission.

Thromboembolic events in the context of COVID-19 are strongly associated with mortality.^3,25,31,33,34^ The etiology of heightened thrombosis remains unclear, although proposed mechanisms have included systemic inflammation, endothelialitis, and activation of the complement system.^35–37^ Increases in a variety of inflammatory pathways, including bradykinin, interleukin-6, C-reactive protein, and growth differentiation factor 15, have been described in COVID-19.^8,9,38–47^ Further, heparin has been shown to block SARS-CoV-2 viral spike protein binding in experimental studies.^48–50^ We postulate that the combination of heparin’s known antithrombotic and anti-inflammatory effects,^6,7^ in addition to viral infectivity attenuation may, at least in part, explain the observed benefit associated with prophylactic anticoagulation.

### Strengths and limitations

While this study had many strengths, including the availability of detailed, longitudinal, electronic health record data on a nationwide cohort of patients hospitalized with COVID-19, rigorous methodology and findings that were robust to sensitivity analyses, we recognize possible limitations. First, due to the observational nature of the study, a degree of uncertainty persists that can only be addressed through randomized trials. Nonetheless, we took several steps to mitigate potential confounding. We comprehensively accounted for chronic and acute health conditions at hospital admission in addition to other potential COVID-19 treatments to achieve balance of these potential confounders between treatment groups. Further, we showed that our results were robust to unmeasured confounding using quantitative bias analysis, which demonstrated that a confounder would need to be strongly associated with receipt of prophylactic anticoagulation and each of the outcomes considered to explain the observed effects. Second, we did not have available a validated algorithm to identify thromboembolic events as an outcome. However, the use of therapeutic anticoagulation may occur as a result of many reported complications of COVID-19 including venous thromboembolism, arterial thromboembolism, cardiac arrhythmia, and disseminated intravascular coagulation.^51–58^ We surmised that an intensification of anticoagulation indicated an adverse change in clinical condition. Third, this study was conducted on Veterans currently receiving care in the VA, who are older and have a higher prevalence of chronic health conditions and risk behaviors than the general US population.^59–61^ However, prior research has established that after adjusting for age, sex, race/ethnicity, region, and rural/urban residence, all of which were accounted for in this study, there is no difference in total disease burden between Veterans and non-Veterans.^61^ Our key finding has also been shown in non-Veteran populations;^11,12^ thus, effects reported in this study are likely generalizable to the wider US population. Fourth, while individuals in VA care represent a diversity of backgrounds, women represented a small proportion of individuals in the sample.

### Summary

We studied a nationwide cohort of patients hospitalized with COVID-19 and found that initiation of prophylactic, heparin-based anticoagulation within 24 hours of admission was associated with a lower risk of 30-day mortality, in-hospital mortality, and initiation of therapeutic-dose anticoagulation likely indicative of a thromboembolic event. Our results provide strong real-world evidence to support guidelines recommending the use of prophylactic anticoagulation as initial therapy for COVID-19 patients upon hospital admission.

## Data Availability

Due to US Department of Veterans Affairs (VA) regulations and our ethics agreements, the analytic data sets used for this study are not permitted to leave the VA firewall without a Data Use Agreement. This limitation is consistent with other studies based on VA data. However, VA data are made freely available to researchers with an approved VA study protocol. For more information, please visit https://www.virec.research.va.gov or contact the VA Information Resource Center at.

## Acknowledgements

The VA Medications, Safety, and Effectiveness Collaboratory Workgroup have vetted the study design and statistical approach prior to study approval.

## Guarantor

CTR and MSF are guarantors.

## Contributorship

Contributions are as follows:

Conceptualization: CTR, JB, ACJ, MSF

Data curation: CTR, CA, FK, MS, YH, MFL, JPT Formal analysis: CTR

Funding acquisition: CTR, DA, ACJ, MSF

Methodology: CTR, JB, LT, WFG, IJD, JPT, SJWE, DA, ACJ, MSF

Project administration: CTR, DA, ACJ, MSF Resources: CTR, DA, ACJ, MSF

Software: CTR, CA, FK, MS, JPT

Supervision: CTR, JB, DA, ACJ, MSF Visualization: CTR

Writing (original draft): CTR, JB

Writing (review & editing): CTR, JB, LT, WFG, CA, FK, MS, EB, JTK, YH, SE, SK, MFL, RAG, PMO, PAH, DAJ, IJD, JPT, SJWE, DA, ACJ, MSF

