## Supplementary Appendix for "Early initiation of prophylactic anticoagulation for prevention of COVID-19 mortality: a nationwide cohort study of hospitalized patients in the United States"

**Table of contents**

Box 1. Agents and doses of prophylactic anticoagulation 2

eFigure 1. Distribution of propensity scores by exposure status before and after weighting 3

eFigure 2. Balance between treatment groups before and after weighting 4

eFigure 3. E-value demonstrating required strength of unmeasured confounder to explain observed associations 5

eTable 1. Sensitivity of IPT-weighted hazard ratios and 95% confidence intervals by capping of propensity score 6

eTable 2. Sensitivity of IPT-weighted hazard ratios and 95% confidence intervals by use of robust variance estimation or stabilized weights 7

eTable 3. Sensitivity analyses expanding exposure ascertainment window from 24 to 48 hours 8

eTable 4. Sensitivity analyses excluding prophylactic direct oral anticoagulants from exposure definition 9

eTable 5. Effect of prophylactic anticoagulation separately by drug 10

STROBE and RECORD statements 11

**Box 1. Agents and doses of prophylactic anticoagulation**

| Type | Drug | Route | Prophylactic dose | Observed in sample |
|---|---|---|---|---|
| Heparin | Heparin | SC | 5000 units b.i.d. or t.i.d. | 1094 (30.2%) |
| LMWH | Enoxaparin | SC | 40 mg q.d. or 30 mg b.i.d. | 2506 (69.1%) |
| LMWH | Fondaparinux | SC | 2.5 mg q.d. | 4 (0.1%) |
| LMWH | Dalteparin | SC | 2500-5000 units q.d. | - |
| DOAC | Apixaban | Oral | 2.5 mg b.i.d. | 21 (0.6%) |
| DOAC | Rivaroxaban | Oral | 10 mg q.d. or 2.5 mg b.i.d. for arterial disease | 2 (0.1%) |
| DOAC | Dabigatran | Oral | 220 mg q.d. | - |

Abbreviations: q.d., once daily; b.i.d., twice daily; t.i.d., thrice daily; SC, subcutaneous; LMWH, low-molecular-weight heparin; DOAC, direct oral anticoagulant

**eFigure 1. Distribution of propensity scores by exposure status before and after weighting**

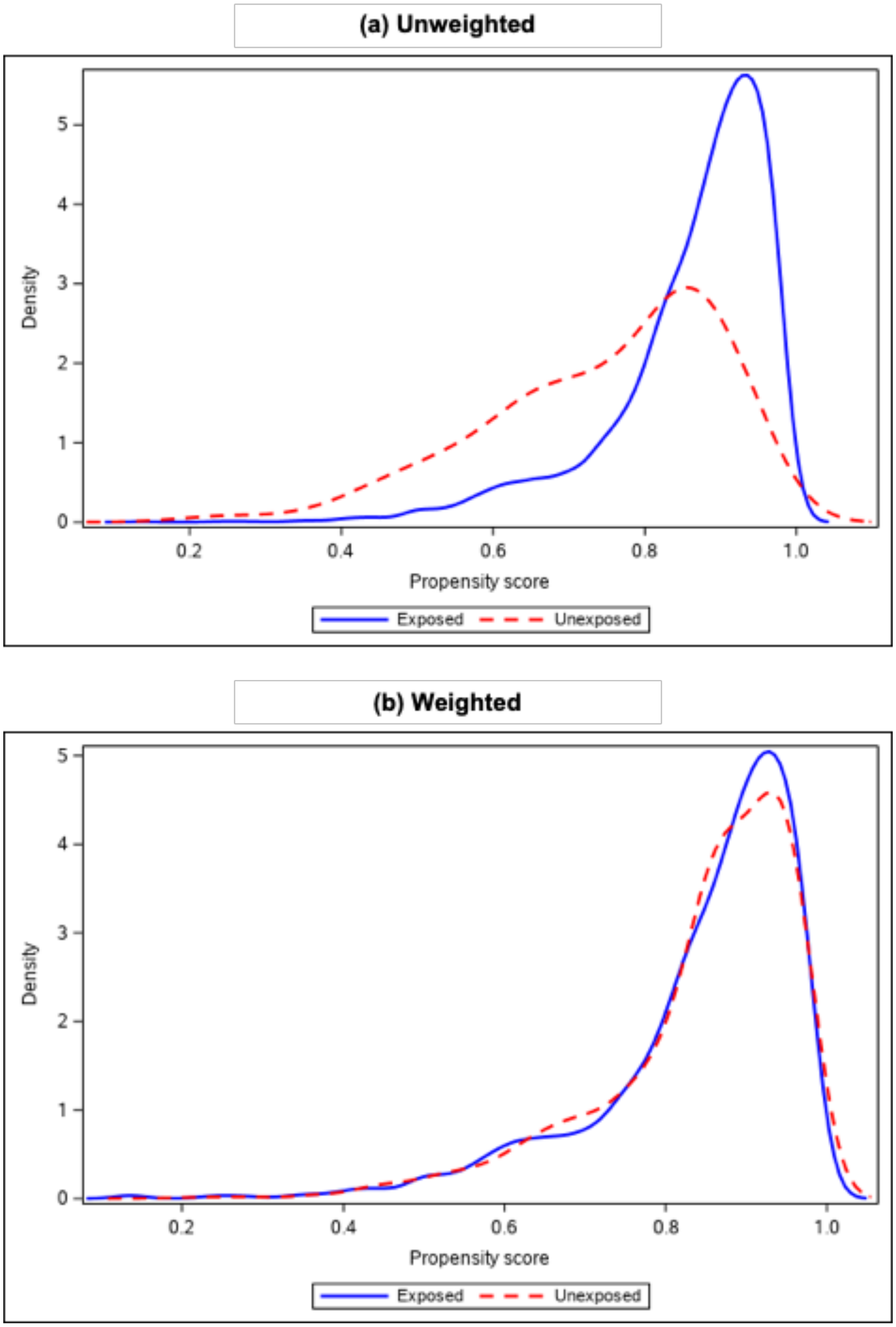

### eFigure 2. Balance between treatment groups before and after weighting

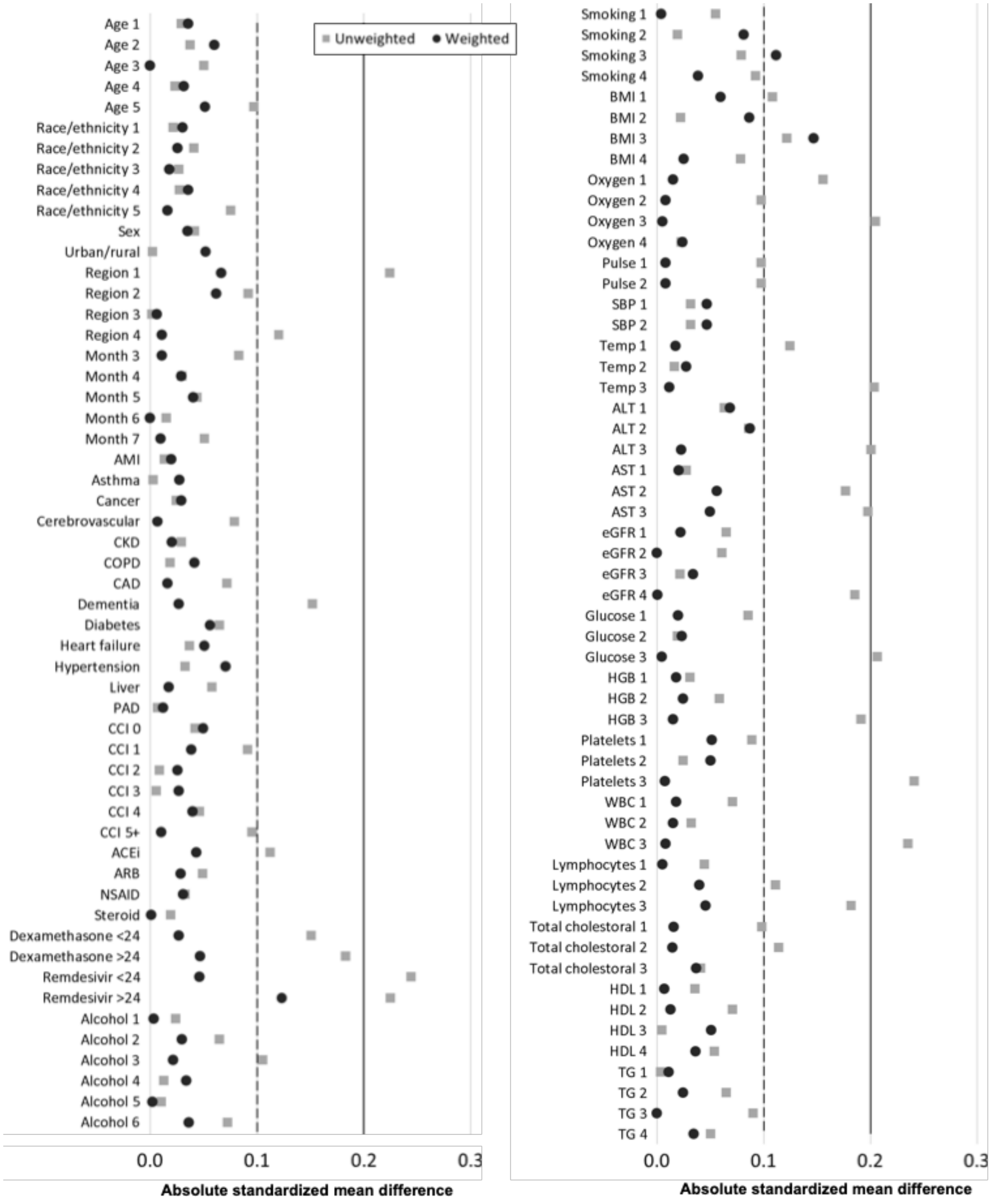


*Note:* Covariates with standardised differences of <0.2 in the weighted sample were considered balanced

### eFigure 3. E-value demonstrating required strength of unmeasured confounder to explain observed associations

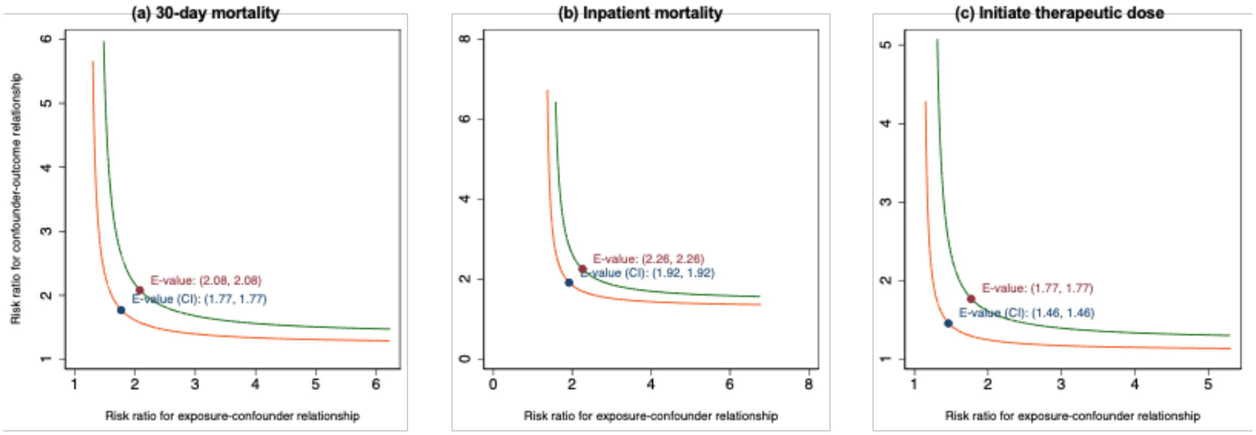


*Note:* Required strength of unmeasured confounder represented in area to the right of curve

### eTable 1. Sensitivity of IPT-weighted hazard ratios and 95% confidence intervals by capping of propensity score

| | | | Cutoff used to cap propensity score | | |
|---|---|---|---|---|---|
| **Outcome** | **N** | **No. events** | **No cap (primary analysis)** | **1st/99th percentile** | **5th/95th percentile** |
| 30-day mortality | | | | | |
| Exposed | 3627 | 513 | 0.73 (0.66-0.81) | 0.76 (0.68-0.84) | 0.79 (0.71-0.88) |
| Unexposed | 670 | 109 | ref | ref | ref |
| Inpatient mortality | | | | | |
| Exposed | 3627 | 418 | 0.69 (0.61-0.77) | 0.71 (0.63-0.80) | 0.74 (0.65-0.83) |
| Unexposed | 670 | 92 | ref | ref | ref |
| Initiate therapeutic anticoagulation | | | | | |
| Exposed | 3627 | 573 | 0.81 (0.73-0.90) | 0.82 (0.74-0.92) | 0.87 (0.78-0.97) |
| Unexposed | 670 | 92 | ref | ref | ref |

Note. Propensity score distributions were capped at 1st/99th percentile and 5th/95th percentile. For example, if an individual had a propensity score lower than the 5th percentile, their propensity score was set to the 5th percentile.

Abbreviations: IPT, inverse probability of treatment

### eTable 2. Sensitivity of IPT-weighted hazard ratios and 95% confidence intervals by use of robust variance estimation or stabilized weights

| | | | Primary analysis (ATE weight) | | Stabilized ATE weight | |
|---|---|---|---|---|---|---|
| **Outcome** | **N** | **No. events** | **Default variance** | **Robust variance** | **Default variance** | **Robust variance** |
| 30-day mortality | | | | | | |
| Exposed | 3627 | 513 | 0.73 (0.66-0.81) | 0.73 (0.55-0.97) | 0.73 (0.60-0.89) | 0.73 (0.55-0.97) |
| Unexposed | 670 | 109 | ref | ref | ref | ref |
| Inpatient mortality | | | | | | |
| Exposed | 3627 | 418 | 0.69 (0.61-0.77) | 0.69 (0.50-0.93) | 0.68 (0.55-0.84) | 0.68 (0.50-0.93) |
| Unexposed | 670 | 92 | ref | ref | ref | ref |
| Initiate therapeutic anticoagulation | | | | | | |
| Exposed | 3627 | 573 | 0.81 (0.73-0.90) | 0.81 (0.61-1.07) | 0.81 (0.66-0.98) | 0.81 (0.61-1.07) |
| Unexposed | 670 | 92 | ref | ref | ref | ref |

Abbreviations: IPT, inverse probability of treatment; ATE, average treatment effect

### eTable 3. Sensitivity analyses expanding exposure ascertainment window from 24 to 48 hours

| | Primary analysis (24 hours) | | | 48 hours | | |
|---|---|---|---|---|---|---|
| Outcome | N | No. events | IPT-weighted HR (95% CI) | N | No. events | IPT-weighted HR (95% CI) |
| 30-day mortality | | | | | | |
| Exposed | 3627 | 513 | 0.73 (0.66-0.81) | 3369 | 492 | 0.81 (0.72-0.91) |
| Unexposed | 670 | 109 | ref | 364 | 66 | ref |
| Inpatient mortality | | | | | | |
| Exposed | 3627 | 418 | 0.69 (0.61-0.77) | 3369 | 413 | 0.88 (0.77-1.00) |
| Unexposed | 670 | 92 | ref | 364 | 54 | ref |
| Initiate therapeutic anticoagulation | | | | | | |
| Exposed | 3627 | 573 | 0.81 (0.73-0.90) | 3369 | 470 | 0.65 (0.58-0.73) |
| Unexposed | 670 | 92 | ref | 364 | 46 | ref |

Note. Lower sample size in the "48 hour" models due to the exclusion of patients who experienced any outcome or discharged during the exposure ascertainment window, which was expanded to 48 hours.

Abbreviations: IPT, inverse probability of treatment; HR, hazard ratio; CI, confidence interval

### eTable 4. Sensitivity analyses excluding prophylactic direct oral anticoagulants from exposure definition

| | Primary analysis | | | Excluding DOACs | | |
|---|---|---|---|---|---|---|
| Outcome | N | No. events | IPT-weighted HR (95% CI) | N | No. events | IPT-weighted HR (95% CI) |
| 30-day mortality | | | | | | |
| Exposed | 3627 | 513 | 0.73 (0.66-0.81) | 3600 | 506 | 0.73 (0.65-0.81) |
| Unexposed | 670 | 109 | ref | 697 | 116 | ref |
| Inpatient mortality | | | | | | |
| Exposed | 3627 | 418 | 0.69 (0.61-0.77) | 3600 | 415 | 0.69 (0.61-0.77) |
| Unexposed | 670 | 92 | ref | 697 | 95 | ref |
| Initiate therapeutic anticoagulation | | | | | | |
| Exposed | 3627 | 573 | 0.81 (0.73-0.90) | 3600 | 563 | 0.79 (0.71-0.88) |
| Unexposed | 670 | 92 | ref | 697 | 102 | ref |

Abbreviations: DOAC, direct oral anticoagulant; IPT, inverse probability of treatment; HR, hazard ratio; CI, confidence interval

### eTable 5. Effect of prophylactic anticoagulation separately by drug

| | Heparin | | | Enoxaparin | | |
|---|---|---|---|---|---|---|
| **Outcome** | **N** | **No. events** | **IPT-weighted HR (95% CI)** | **N** | **No. events** | **IPT-weighted HR (95% CI)** |
| 30-day mortality | | | | | | |
| Exposed | 1094 | 230 | 0.73 (0.64-0.84) | 2506 | 276 | 0.78 (0.68-0.89) |
| Unexposed | 670 | 109 | ref | 670 | 109 | ref |
| Inpatient mortality | | | | | | |
| Exposed | 1094 | 196 | 0.69 (0.60-0.80) | 2506 | 219 | 0.72 (0.62-0.84) |
| Unexposed | 670 | 92 | ref | 670 | 92 | ref |
| Initiate therapeutic anticoagulation | | | | | | |
| Exposed | 1094 | 171 | 0.96 (0.81-1.13) | 2506 | 392 | 0.79 (0.70-0.89) |
| Unexposed | 670 | 92 | ref | 670 | 92 | ref |

Abbreviations: IPT, inverse probability of treatment; HR, hazard ratio; CI, confidence interval

### STROBE and RECORD statements

| | Item No. | STROBE items | Location in manuscript where items are reported | RECORD items | Location in manuscript where items are reported |
|---|---|---|---|---|---|
| **Title and abstract** | | | | | |
| | 1 | (a) Indicate the study's design with a commonly used term in the title or the abstract (b) Provide in the abstract an informative and balanced summary of what was done and what was found | (a) Title & Abstract<br><br>(b) Abstract | RECORD 1.1: The type of data used should be specified in the title or abstract. When possible, the name of the databases used should be included.<br><br>RECORD 1.2: If applicable, the geographic region and timeframe within which the study took place should be reported in the title or abstract.<br><br>RECORD 1.3: If linkage between databases was conducted for the study, this should be clearly stated in the title or abstract. | 1.1: Abstract<br><br>1.2: Abstract – Setting & Participants<br><br>1.3: N/A |
| **Introduction** | | | | | |
| Background rationale | 2 | Explain the scientific background and rationale for the investigation being reported | Background (Para 1) | | |
| Objectives | 3 | State specific objectives, including any prespecified hypotheses | Background (End of Para 2) | | |
| **Methods** | | | | | |
| Study Design | 4 | Present key elements of study design early in the paper | Methods (Study design and population) | | |
| Setting | 5 | Describe the setting, locations, and relevant dates, including periods of recruitment, exposure, follow-up, and data collection | Methods (Study design and population; Exposure, outcomes and follow-up) | | |
| Participants | 6 | *(a) Cohort study* - Give the eligibility criteria, and the sources and methods of selection of participants. Describe methods of follow-up *Case-control study* - Give the eligibility criteria, and the sources and methods of case ascertainment and control selection. Give the rationale for the choice of cases and controls | (a) Methods (Study design and population) | RECORD 6.1: The methods of study population selection (such as codes or algorithms used to identify subjects) should be listed in detail. If this is not possible, an explanation should be provided.<br><br>RECORD 6.2: Any validation studies of the codes or algorithms used to select the population should be referenced. If | 6.1: Methods (Study design and population)<br><br>6.2: N/A<br><br>6.3: N/A |

| | | | | | |
|---|---|---|---|---|---|
| | | *Cross-sectional study* - Give the eligibility criteria, and the sources and methods of selection of participants<br><br>*(b) Cohort study* - For matched studies, give matching criteria and number of exposed and unexposed<br>*Case-control study* - For matched studies, give matching criteria and the number of controls per case | | validation was conducted for this study and not published elsewhere, detailed methods and results should be provided.<br><br>RECORD 6.3: If the study involved linkage of databases, consider use of a flow diagram or other graphical display to demonstrate the data linkage process, including the number of individuals with linked data at each stage. | |
| Variables | 7 | Clearly define all outcomes, exposures, predictors, potential confounders, and effect modifiers. Give diagnostic criteria, if applicable. | Methods (Forms and doses of anticoagulation; Exposure, outcomes, and follow-up, Covariates; Figure 1) | RECORD 7.1: A complete list of codes and algorithms used to classify exposures, outcomes, confounders, and effect modifiers should be provided. If these cannot be reported, an explanation should be provided. | 7.1: Methods (Forms and doses of anticoagulation; Exposure, outcomes, and follow-up, Covariates; Figure 1) |
| Data sources/ measurement | 8 | For each variable of interest, give sources of data and details of methods of assessment (measurement).<br>Describe comparability of assessment methods if there is more than one group | Methods (Forms and doses of anticoagulation; Exposure, outcomes, and follow-up, Covariates; Figure 1) | | |
| Bias | 9 | Describe any efforts to address potential sources of bias | Methods (Propensity score model; Statistical methods; Sensitivity analyses) | | |
| Study size | 10 | Explain how the study size was arrived at | Methods (Study design and population; Figure 2) | | |
| Quantitative variables | 11 | Explain how quantitative variables were handled in the analyses. If applicable, describe which groupings were chosen, and why | Methods (Propensity score model; Statistical methods) | | |
| Statistical methods | 12 | (a) Describe all statistical methods, including those used to control for confounding<br>(b) Describe any methods used to examine subgroups and interactions | (a-c) Methods (Covariates; Propensity score model; Statistical methods, Sensitivity analyses)<br><br>(d) N/A | | |

| | | | | | |
|---|---|---|---|---|---|
| | | (c) Explain how missing data were addressed<br>(d) *Cohort study* - If applicable, explain how loss to follow-up was addressed<br>*Case-control study* - If applicable, explain how matching of cases and controls was addressed<br>*Cross-sectional study* - If applicable, describe analytical methods taking account of sampling strategy<br>(e) Describe any sensitivity analyses | (e) Methods (Sensitivity analyses) | | |
| Data access and cleaning methods | | .. | | RECORD 12.1: Authors should describe the extent to which the investigators had access to the database population used to create the study population.<br><br>RECORD 12.2: Authors should provide information on the data cleaning methods used in the study. | 12.1: Methods (Study design and population; Covariates; Ethics; Data sharing; Funding; Contributorship)<br><br>12.2: Methods (Covariates) |
| Linkage | | .. | | RECORD 12.3: State whether the study included person-level, institutional-level, or other data linkage across two or more databases. The methods of linkage and methods of linkage quality evaluation should be provided. | N/A |
| **Results** | | | | | |
| Participants | 13 | (a) Report the numbers of individuals at each stage of the study (*e.g.*, numbers potentially eligible, examined for eligibility, confirmed eligible, included in the study, completing follow-up, and analysed)<br>(b) Give reasons for non-participation at each stage.<br>(c) Consider use of a flow diagram | (a-c) Results (Patient characteristics, Figure 2) | RECORD 13.1: Describe in detail the selection of the persons included in the study (*i.e.,* study population selection) including filtering based on data quality, data availability and linkage. The selection of included persons can be described in the text and/or by means of the study flow diagram. | 13.1: Results (Patient characteristics, Figure 2) |
| Descriptive data | 14 | (a) Give characteristics of study participants (*e.g.*, demographic, clinical, social) and information on | (a) Results (Patient characteristics, Table 1)<br><br>(b) Table 1 | | |

| | | | | | |
|---|---|---|---|---|---|
| | | exposures and potential confounders<br>(b) Indicate the number of participants with missing data for each variable of interest<br>(c) *Cohort study* - summarise follow-up time (*e.g.*, average and total amount) | (c) Results (Patient characteristics) | | |
| Outcome data | 15 | *Cohort study* - Report numbers of outcome events or summary measures over time<br>*Case-control study* - Report numbers in each exposure category, or summary measures of exposure<br>*Cross-sectional study* - Report numbers of outcome events or summary measures | Results (Absolute and relative risks) | | |
| Main results | 16 | (a) Give unadjusted estimates and, if applicable, confounder-adjusted estimates and their precision (e.g., 95% confidence interval). Make clear which confounders were adjusted for and why they were included<br>(b) Report category boundaries when continuous variables were categorized<br>(c) If relevant, consider translating estimates of relative risk into absolute risk for a meaningful time period | (a) Results (Absolute and relative risks; Figure 3; Table 2)<br><br>(b) Results (Table 1)<br><br>(c) Results (Absolute and relative risks; Table 2) | | |
| Other analyses | 17 | Report other analyses done—e.g., analyses of subgroups and interactions, and sensitivity analyses | Results (Sensitivity analyses; Supplementary Appendix) | | |
| **Discussion** | | | | | |
| Key results | 18 | Summarise key results with reference to study objectives | Discussion (Key findings) | | |
| Limitations | 19 | Discuss limitations of the study, taking into account sources of potential bias or imprecision. Discuss both direction and magnitude of any potential bias | Discussion (Strengths and limitations) | RECORD 19.1: Discuss the implications of using data that were not created or collected to answer the specific research question(s). Include discussion of misclassification bias, unmeasured confounding, missing data, and changing | 19.1 Discussion (Comparison with other evidence; Strengths and limitations) |

| | | | | | |
|---|---|---|---|---|---|
| | | | | eligibility over time, as they pertain to the study being reported. | |
| Interpretation | 20 | Give a cautious overall interpretation of results considering objectives, limitations, multiplicity of analyses, results from similar studies, and other relevant evidence | Discussion (Throughout) | | |
| Generalisability | 21 | Discuss the generalisability (external validity) of the study results | Discussion (Strengths and limitations) | | |
| **Other Information** | | | | | |
| Funding | 22 | Give the source of funding and the role of the funders for the present study and, if applicable, for the original study on which the present article is based | Funding | | |
| Accessibility of protocol, raw data, and programming code | | .. | | RECORD 22.1: Authors should provide information on how to access any supplemental information such as the study protocol, raw data, or programming code. | Data sharing |
